# Identification of Vulnerable Populations and Areas at Higher Risk of COVID-19 Related Mortality in the U.S.

**DOI:** 10.1101/2020.07.11.20151563

**Authors:** Esteban Correa-Agudelo, Tesfaye B. Mersha, Andrés Hernández, Adam J. Branscum, Neil J. MacKinnon, Diego F. Cuadros

## Abstract

**Background:** The role of health-related disparities including sociodemographic, environmental, and critical care capacity in the COVID-19 pandemic are poorly understood. In the present study, we characterized vulnerable populations located in areas at higher risk of COVID-19 related mortality and low critical healthcare capacity in the U.S.

**Methods:** Using Bayesian multilevel analysis and small area disease risk mapping, we assessed the spatial variation of COVID-19 related mortality risk for the U.S. in relation with healthcare disparities including race, ethnicity, poverty, air quality, and critical healthcare capacity.

**Results:** Overall, highly populated, regional air hub areas, and minorities had an increased risk of COVID-19 related mortality. We found that with an increase of only 1 ug/m3 in long term PM2.5 exposure, the COVID-19 mortality rate increased by 13%. Counties with major air hubs had 18% increase in COVID-19 related death compared to counties with no airport connectivity. Sixty-eight percent of the counties with high COVID-19 related mortality risk were also counties with lower critical care capacity than national average. These counties were primary located at the North- and South-Eastern regions of the country.

**Conclusion:** The existing disparity in health and environmental risk factors that exacerbate the COVID-19 related mortality, along with the regional healthcare capacity, determine the vulnerability of populations to COVID-19 related mortality. The results from this study can be used to guide the development of strategies for the identification and targeting preventive strategies in vulnerable populations with a higher proportion of minority groups living in areas with poor air quality and low healthcare capacity.

**KEY POINTS:** *Question:* What are the sociodemographic and environmental drivers of the heterogeneous distribution of the COVID-19 related mortality in the U.S., and what are the vulnerable areas at higher risk of COVID-19 related mortality and low critical healthcare capacity?

*Findings:* Higher proportions of African American and Latino populations, as well as high levels of air pollution and airport connectivity were linked to higher risk of COVID-19 related mortality. Over 68% of the counties with high COVID-19 related mortality risk were also counties with lower critical care capacity than national average.

*Meaning:* In a time-limited response, the identification and targeting prevention efforts should focus in vulnerable populations located in high risk areas in which sociodemographic and environmental factors are exacerbating the burden of COVID-19 related deaths.

## INTRODUCTION

Early Coronavirus Disease 2019 (COVID-19) data from Europe and Asia suggested unprecedented contagious and death rates of the pandemic. In late March, the United States (U.S.) exhibited the fastest growing curve in terms of deaths across developed countries, with 98,768 as of May 26^th^. In terms of disease characterization, several countries along with U.S. have reported higher mortality rates (MR) for the older population with concomitant comorbidities including chronic lower respiratory diseases, diabetes, hypertension and ischemic diseases among others^1^. Preliminary studies have started to create baseline population characteristics of COVID-19 related deaths^2^ and public health researchers have started to project next steps in terms of the disease control strategies and healthcare resource allocations and demand^3,4^. However, the role of geospatial disparities, including sociodemographic and environmental exposures, and critical care capacity for the future of the pandemic are poorly understood. Identifying which population groups and areas who have a higher risk of COVID-19 mortality based on underlying health disparities and low critical healthcare capacity is a logical step to develop more effective strategies for mitigating the risk where more susceptible populations reside. Under the light of these considerations, this study aims to: a) assess the sociodemographic and environmental drivers of COVID-19 related deaths, and b) spatially identify vulnerable areas at higher risk of COVID-19 mortality but with low healthcare capacity. We hypothesized that COVID-19 related mortality will significantly affect counties with predominant minority groups, poor air quality and low critical healthcare capacity in the U.S.

## RESEARCH DESIGN AND METHODS

### Study area and data sources

The U.S. COVID-19 data were obtained from the Johns Hopkins University dataset^5^ for 3,009 counties from January 22, 2020, to May 26, 2020 including 49 states. Since we were interested in identifying general patters of COVID-related mortality for the entire U.S., we excluded data from New York state, which has experienced an unusual intensive COVID-19 outbreak, behaving as a hotspot with about 5% of worldwide cases^6^. Sociodemographic data were derived from recent American Community Survey 2014-2018 5-Year Estimates (ACS)^7^, and the Center for Disease Control (CDC) Social Vulnerability Index^8^. Due to the strong association of COVID-19 with underlying health problems, county-level comorbidities, including chronic lower respiratory disease (CLRD), diabetes mellitus, hypertensive diseases (HTA), and ischemic heart disease were obtained between 2010 to 2018^9^. Similarly, air pollution was assessed using the Surface annual PM_2.5_ satellite images from 2000 to 2018^10^.

### Study variables

The primary outcome of interest was COVID-19 related deaths. County-level cumulative number of deaths up to May 26, 2020 were included as the health outcome measure. All covariates were selected according to an evidence synthesis process of preliminary reported results^2,11-15^, and results were aggregated and reported at county level. A directed acyclic graph (DAG) was built to infer causal effects to the observational data. Next, we removed open paths, check for colliders and overcontrol in the implied graph (Figure S1)^16^. The socioeconomic and demographic variables included percentage estimated for total population by age-groups (Under 25, 25-34, 35-44, 45-59, 60-74, and over 75), percentage estimated for total population of self-identified as White, African American, and Hispanic or Latino ethnicity by county according to the Census Bureau definition^17^, and percentage estimated for persons below poverty according to the CDC’s vulnerability index^8^.

For county-level underlying cause of death, we selected four chronic conditions including: CLRD, diabetes mellitus, HTA, and ischemic heart disease MR per 100,000 people^15^. To evaluate the link between environmental exposures and COVID-19 related mortality, we used 2000 to 2018 annual images of ground-level fine particle matter (PM_2.5_) over North America^10^. These calibrated images are estimated at a 0.01° × 0.01° grid resolution combining satellite and monitoring stations data sources using a Geographically Weighted Regression (GWR). Since the unit of analysis for this ecological study is the county, we aggregated PM_2.5_ data at a county-level resolution. Then, we computed long-term exposure by temporally averaging PM_2.5_ between 2010 to 2018 within each county. We also calculated regional air hub and road connectivity index for each county to examine the association of deaths and county-level airport hubs and main roads. We generated four levels of connectivity index as following: counties with an airport with more than 50,000 passengers per year (Has an airport), counties next to a county with an airport (Next to airport), counties crossed by a main road (Crossed by a highway), and counties not surrounded with a county with an airport and not being crossed by a main road (No airport/highway). A more detailed information of covariate description is included in Supplementary Materials (see Appendix A). This study follows the guidelines of the Strengthening the Reporting of Observational Studies in Epidemiology (STROBE)^18^.

### Multivariate analyses of risk factors for COVID-19 related death

In this study, a Bayesian multilevel analysis was used to assess the risk of COVID-19 related death per county adjusting for covariates. We included the cumulative number of deaths for the observed variable and the projection of expected deaths using ACS population as the regression offset. Previous studies showed strong association between age and COVID-19 death counts, so, as a result, we adjusted all models with age-group population distributions. We included a random intercept at state-level to assess group effects. Normal and Half Cauchy weak informative priors^19^, four Monte Carlo Markov Chains (MCMC), and 4,000 iterations with No-U-Turn Sampler (NUTS)^20^ were used to fit the model. All numeric covariates where centered for easily interpretation on national average.

### COVID-19 disease mapping

We generated small area disease risk map after adding state-level random intercepts. Small area risk estimates were generated by computing the crude mortality rate (CMR) for each county. The CMR was obtained as the ratio of observed (*Y*_*i*_) to the expected disease counts (*E*_*i*_): *CMR*_*i*_=*Y*_*i*_/*E*_*i*_, where the expected counts represented the total number of COVID-19 related deaths based on the population of the specific area (ACS county population). A Poisson distribution was used to avoid extreme values due to areas with small populations. Counties with relative risk (RR) equal to one have the same risk as expected based on the total population of the county. Counties with RR less than one indicates lower relative risk, and greater than one is an evidence of a higher mortality risk than average. Quantile population classification was used to identify the ten highest COVID-19 mortality risk areas in highly populated counties (4^th^ quartile). A bivariate map combining COVID-19 related mortality risk and number of intensive care units (ICU) beds was generated to identify key vulnerable areas with low critical healthcare capacity. ICU beds per 100,000 people were included as an index of critical healthcare capacity of each county^21^. Both variables were classified with a Tertile scheme as follows: COVID-19 related RR (0-1 lower risk, 1-3, medium risk, 3 > high risk), ICU beds per 100,000 inhabitants (< 28.4 low availability, 28.4-100 medium availability, > 100 high availability). The R language including brms, INLA, SpatialEpi, and raster packages were used to implement all models and maps^22-25^. A more detailed information including equations is included in Supplementary Materials (see Appendix B).

## RESULTS

### General results

Table 1 shows descriptive statistics of COVID-19 deaths in the 49 states included in the study. The total number of deaths reported were 68,288, corresponding to 5.3% of the 1,300,169 COVID-19 confirmed cases (excluding the state of NY). Highest cumulative death counts were found in Cook County, IL (3,354 deaths), Wayne County, MI (2,368 deaths), and Los Angeles County, CA (2,45 deaths) respectively. Of the 3,009 counties included in the study, 1,703 had at least one confirmed COVID-19 death, and 570 counties had no valid information for all covariates, leaving a sample of 2,439 counties (excluding the state of NY). The overall percent estimate of poverty was 15.6 (standard deviation [SD] 6.5) for the entire country. White population had an average proportion of 83.0% (SD 16.7), African American 9.1% (SD 14.6) and Latino 9.3% (SD 13.9). The national average PM_2.5_ exposure was 8.0 µg/m (SD 2.4). For the connectivity index, 220 counties had an airport with more than 50,000 passengers per year, 619 counties had a highway or main road, and 1,194 counties are categorized as low transportation connectivity. The overall ICU beds capacity was 28.4 per 100,000 (SD 34.6).

**Table 1.** Baseline characteristics.

| **County-level covariates** |  |  |
| --- | --- | --- |
| COVID-19, n |  |  |
| Confirmed cases | 1,300,169 |  |
| Confirmed deaths | 68,288 |  |
| Sociodemographic, mean (SD) |  |  |
| Age |  |  |
| Under 25 | 31.2 | (4.8) |
| 25-34 | 11.8 | (2.3) |
| 35-44 | 11.6 | (1.6) |
| 45-59 | 20.2 | (2.2) |
| 60-74 | 17.4 | (3.7) |
| 75+ | 7.9 | (2.4) |
| Estimated % of population in poverty | 15.6 | (6.5) |
| Estimated % of white population | 83.0 | (16.9) |
| Estimated % of African American population | 9.1 | (14.6) |
| Estimated % of Latino population | 9.3 | (13.9) |
| Crude Mortality Rates, mean (SD) |  |  |
| Chronic Lower Respiratory Disease | 70.2 | (26.1) |
| Diabetes Mellitus | 33.7 | (14.7) |
| Hypertension | 27.2 | (17.0) |
| Ischemic Heart Disease | 151.0 | (57.5) |
| Environment, mean (SD) |  |  |
| Long-term exposure to PM2.5 $\mu\text{g}/\text{m}^3$ | 8.0 | (2.4) |
| Connectivity risk, n |  |  |
| No airport/highway | 1194 |  |
| Crossed by a highway | 619 |  |
| Next to Airport | 1013 |  |
| Has an airport | 220 |  |

COVID-19 related MR per 100,0000 people revealed higher average MR in the 4^th^ quartile of counties with PM_2.5_ greater than 10.1 µg/m (21.4 per 100,000) and counties with an airport (17.9 per 100,000), For minority groups, counties with high percentage of African American (25.6 per 100,000) and Latino population (14.8 per 100,000) showed higher COVID-19 related MR (Figure S2).

### Multivariate analyses of risk factors for COVID-19 related death

Figure 1 illustrates the RR for COVID-19 at the state-level. Ten of the 49 states had a risk higher than average (AZ, CO, CT, IN, LA, MA, MI, MS, NJ, and PA). Notably, three of four Northeastern states had the highest RR (excluding NY), CT (RR=8.16, credible interval [CI]: 3.60 -18.73) MA (RR=9.35, CI: 4.76 - 18.54), and NJ (RR=6.69, CI: 3.71 - 12.16). IN and MI (Midwest), LA and MS (South), AZ and CO (West) hold a higher RR than average. Conversely, 11 of the 49 states from Midwest (MO, and SD), North-East (RI), South (AR, TN, TX), and West (AK, CA, HI, UT, and WY) had a RR lower than the national average. Map in Figure 2A illustrates the RR by county and Table 2 shows the ten highest COVID-19 mortality areas in highly populated counties (4^th^ quartile). CT, GA, MI, NJ, and NM top the list with RR at least five-fold higher than average. Also, these counties exhibited on average higher proportions of population in poverty (17.5%), African American (22.4%) and Latino (21.7%) populations compared to the national averages of 15.6%, 9.1%, and 9.3%, respectively. Likewise, nine of these ten counties had a long-term PM_2.5_ exposure of at least 2.6 µg/m above the national average (8.0 µg/m). Eight of these ten counties had an airport or were next to a county with an airport, and five out of ten have lower ICU beds availability than national average of 28.4 ICU beds per 100,000 inhabitants.

**Table 2.** Ten highest COVID-19 mortality risk areas in highly populated counties (4^th^ quartile).

| State | County | Observed counts | Expected counts | Connectivity | PM25 (u/gml) | Uninsured (%) | Poverty (%) | ICU per 100,000 | White race (%) | African American race (%) | Latino ethnicity (%) | RR CI: [2.5%, 97.5%] |
| --- | --- | --- | --- | --- | --- | --- | --- | --- | --- | --- | --- | --- |
| NJ | Essex | 1608 | 179 | Has an airport | 11.2 | 12.3 | 16.4 | 28.5 | 42.1 | 39.8 | 22.7 | 8.99 [8.55, 9.43] |
| NJ | Union | 1030 | 125 | Next to airport | 11.4 | 11.8 | 9.8 | 13.9 | 56.2 | 21.2 | 31.1 | 8.24 [7.75, 8.75] |
| NJ | Passaic | 892 | 113 | Next to Airport | 9.6 | 12.4 | 16.7 | 10.5 | 62.2 | 11.4 | 40.9 | 7.89 [7.38, 8.42] |
| NJ | Hudson | 1143 | 151 | Next to Airport | 12.3 | 14.1 | 16.3 | 13.3 | 55.1 | 12.4 | 43.2 | 7.57 [7.14, 8.02] |
| NJ | Bergen | 1528 | 209 | Next to Airport | 11.3 | 8.3 | 7.0 | 13.1 | 71.4 | 6.0 | 19.4 | 7.31 [6.95, 7.68] |
| GA | Dougherty | 142 | 21 | No airport/<br>highway | 10.8 | 17 | 29.4 | 54.9 | 27 | 69.0 | 2.7 | 6.73 [5.66, 7.87] |
| MI | Wayne | 2368 | 397 | Has an airport | 11.6 | 7.2 | 23.1 | 38.4 | 53.2 | 38.9 | 5.9 | 5.96 [5.73, 6.21] |
| CT | Hartford | 1193 | 201 | Has an airport | 8.5 | 4.4 | 11.1 | 53.2 | 71.5 | 13.6 | 17.6 | 5.94 [5.6, 6.28] |
| NM | McKinley | 95 | 16 | Crossed by a highway | 3.0 | 20.9 | 36 | 35.7 | 15 | 0.7 | 14.3 | 5.83 [4.71, 7.06] |
| CT | Fairfield | 1231 | 213 | Next to Airport | 9.0 | 8.7 | 8.8 | 18.3 | 73.2 | 11.4 | 19.3 | 5.78 [5.46, 6.11] |

**Figure 1.**
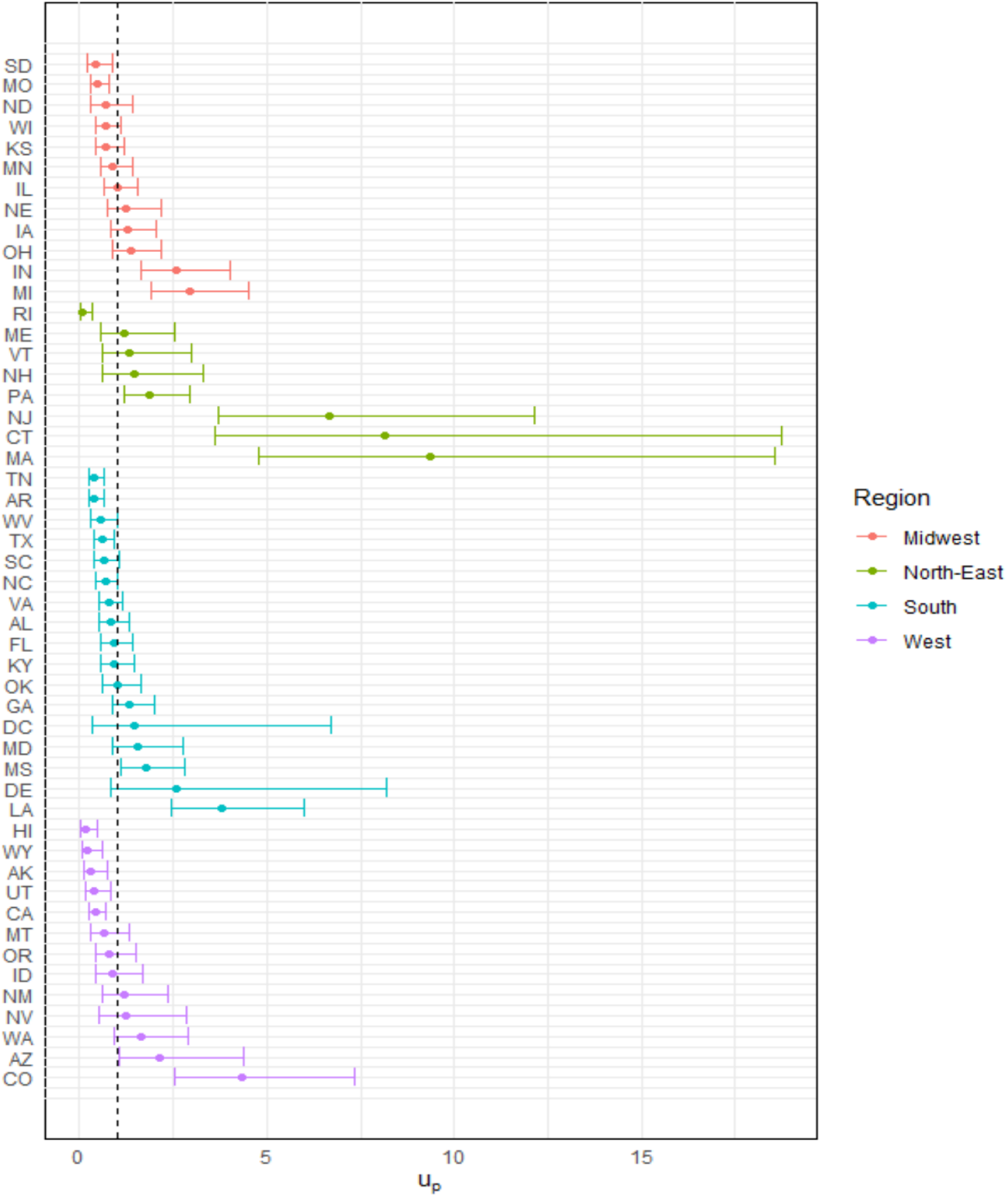
U.S. Relative Risk for COVID-19 by State

**Figure 2.**
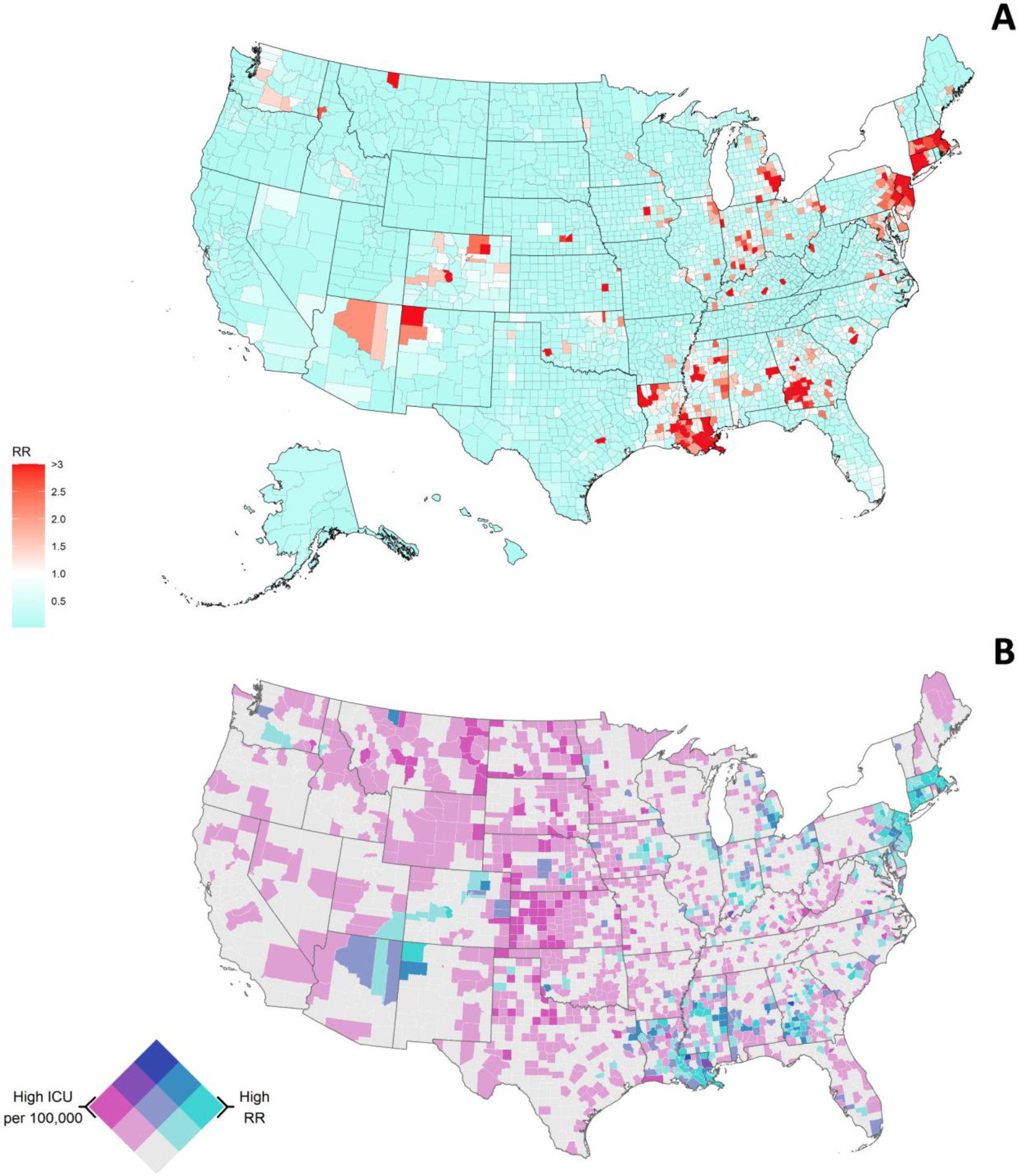
U.S. Relative Risk for COVID-19 by County, mean=0.53 (range is 0.01-14.37) (**A**). U.S. COVID-19 related relative risk (RR) of Death and ICU availability per 100,000, (without NY). Dark purple indicates counties with high ICU availability and low mortality risk whereas areas in darker green-blue indicate counties with high mortality risk but low ICU availability. Both variables were classified with a Tertile scheme as follows: COVID-19 related RR (0-1 lower risk, 1-3 medium risk, 3 > high risk), ICU beds per 100,000 (< 28.4 low availability, 28.4-100 medium availability, > 100 high availability) (**B**)

Table 3 summarizes the RR and credible intervals from the adjusted model for the overall association between COVID-19 related deaths and the covariates in all counties included in the study. For sociodemographic risk factors, the proportion of people living in poverty in the county (mean [*μ*] = 1.01, credible interval [CI]: 1.01-1.02), and the proportion of Latino population if infected with COVID-19 (*μ* = 1.01, 95% CI: 1.01-1.02) were factors associated with higher risk of COVID-19 related death, whereas countries with high proportion of White population had lower risk of COVID-related death (*μ* = 0.97, CI: 0.97-0.97). We found no statistically significant association between the proportion of comorbidities and the risk of COVID-19 related death at county level, except for a negative association with CLRD. For the long-term exposure to air pollution, we found that one additional unit of PM_2.5_ (1.0 µg/m) increased the risk of COVID-related death by 13% (*μ* = 1.13, CI: 1.11-1.14). Lastly, counties with an airport and near to airports had a higher risk of COVID-19 related death compared to counties with low transportation connectivity, with an 18% (*μ* = 1.18, CI: 1.12-1.24), and 16% (*μ* = 1.16, CI: 1.10-1.22) higher mortality risk respectively. A more detailed information including model performance and unadjusted models is included in Supplementary Materials (see Appendix B).

**Table 3.**
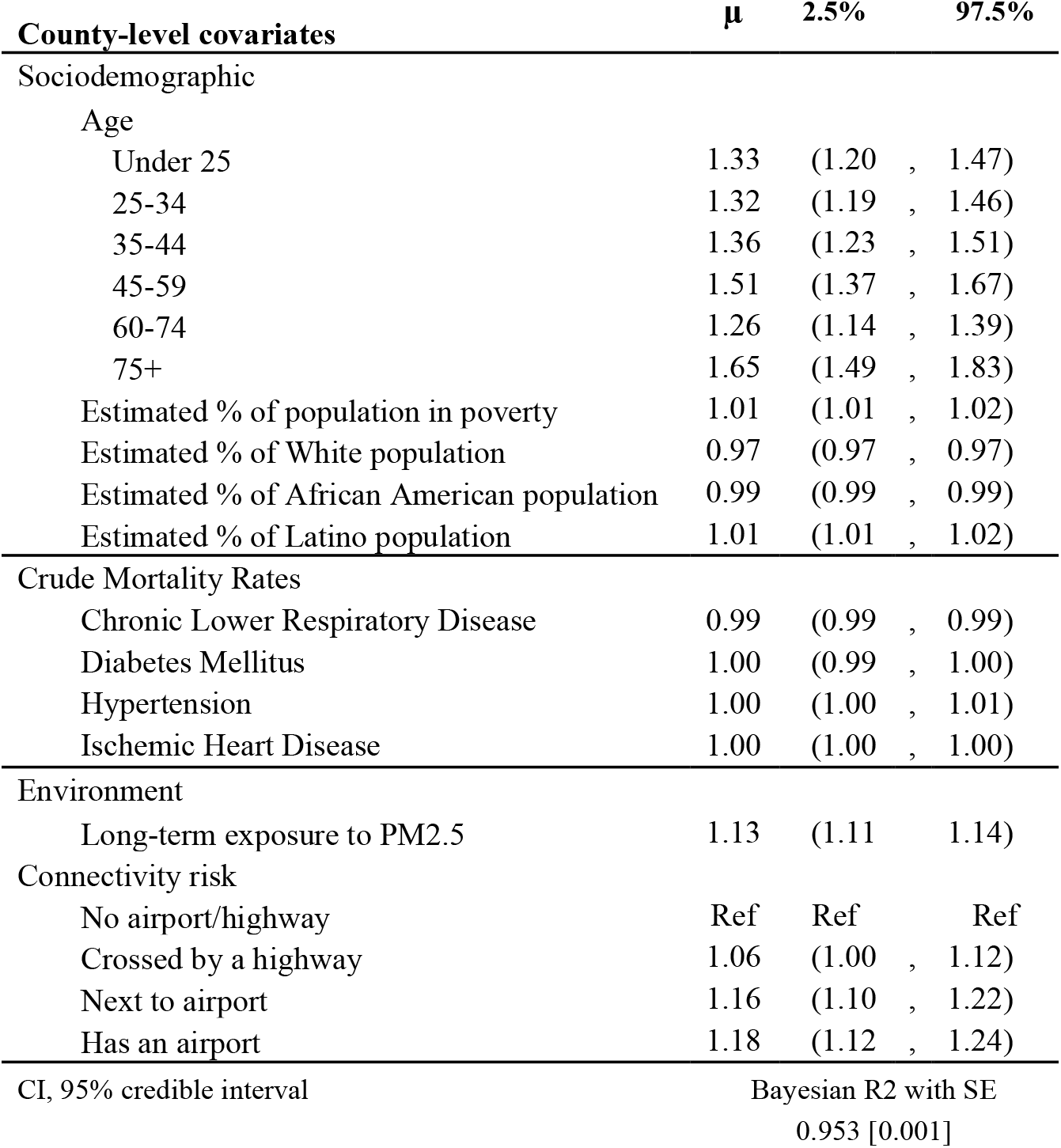
Adjusted county-level predictors of COVID19 deaths

### COVID-19 disease mapping

Overall, we found that 396 counties from 39 states had higher risk of COVID-19 related mortality (RR > 1) (Figure 2A and Table 2). Five states had at least 21 counties with high mortality risk including GA (43), IN (28), LA (42), MS (31), and NJ (21). Figure 2B illustrates the bivariate map of the COVID-19 related mortality risk and ICU beds availability for all counties in the conterminous U.S. We observed 105 counties with high mortality risk, and 71 of these counties with low ICU availability. About 46% of these counties were in GA (10), LA (11), and NJ (12). The map also showed counties with high COVID-19 related mortality risk - high ICU beds per 100,000 such as East Feliciana parishes (LA), and Upson County (GA). Also, areas with low risk of COVID-19 related mortality risk but high ICU availability were observed in KS and ND, as well as several counties with low mortality risk and acceptable ICU capacity at the moment of this analysis. A more detailed information including additional maps and the complete mortality risk list for all states can be found in Supplementary Materials (see Appendix B and C).

## DISCUSSION

This study provides state and county-level characterization of the COVID-19 related mortality risk including sociodemographic and socio-environmental factors across the U.S. Also, our study assessed the spatial link between COVID-19 related mortality risk and the current critical healthcare capacity across the U.S. Overall, we identified highly populated and polluted areas, regional air hub areas, and minorities with an increased COVID-19 related risk of death. The ten most populated counties with the highest mortality risk showed a five-fold higher than national average with higher proportions of African Americans and Latino groups residing in these counties. Moreover, our spatial analysis showed that 68% of the counties with high COVID-19 related mortality risk were also counties with a lower capacity of ICU beds than national average.

We found that the ten highest COVID-19 mortality areas in highly populated counties (4^th^ quartile) showed at least five-fold higher mortality risk than national average. Noteworthy, these counties exhibited on average higher proportions population in poverty, African American and Latino populations compared to their national average. States and counties with historically higher proportion of African American and Latino population such as LA, and NJ were at greater COVID-19 related mortality risk than other states. These demographic disparities in terms of COVID-19 related mortality have been recognized in preliminary results of several major cities in other countries including London^11,26,27^. Moreover, non-pharmaceutical interventions (school closing, physical distancing, lockdowns and additional sanitation), which are the only interventions available to tackle the pandemic,^28^ are difficult to implement in these groups. As a result, the effectiveness and benefit of these non-pharmaceutical interventions can be diluted by the work activities that involve person to person interaction and are more common in these low-income groups exposing them to a higher risk of infection and thus, higher mortality risk.

Air pollution was positively associated with higher COVID-19 related mortality risk, and the top ten counties with the highest mortality risk exhibited higher levels of PM_2.5_ exposure compared to the national average. Air pollution is one of the leading risk factors for respiratory related death globally^27^, and this factor could be playing a key role in exacerbating the numbers of COVID-19 related deaths in highly polluted areas. Air pollution has an indirect impact on most of the organs and systems of human body and indirectly comorbidities. Although we did not find any significant association between COVID-19 and comorbidities at county level, air pollution has been identified as contributing factor for many respiratory diseases like chronic obstructive pulmonary disease (COPD)^26,29^, asthma^29-34^, and lung cancer^35-38^, which are concomitant comorbidities that reported strong association of COVID-19 related deaths at individual level^39^. The health effects of air pollution depend on the components and sources of pollutants, which can vary among counties, seasons, and times. Initial evidence of incidence and mortality with comorbidities have been reported in Italy with strong regional differences between the northern and southern region^3,13^. Although we found a strong positive association between air pollution and the risk of COVID-19 related death, the role of long-term exposure to poor air quality in the actual numbers of COVID-19 related deaths in the U.S. is still not well understood, and thus more studies are needed including major U.S. cities taking into account long-term exposure of outdoor and indoor pollution. Furthermore, our results suggest that counties with airports have higher COVID-19 related mortality risk than those with less connectivity. The high connectivity and travelers in these counties generated by airports can produce a high influx of locally imported infections that boost the local transmission of the virus in the county and consequently the number of COVID-19 related deaths.

According to our spatial analysis, 396 counties (16.2% of the total number of counties included in the study) from 39 states had higher risk of COVID-19 related mortality than the national average. About 56% (221 out of 396) of these counties where located in only eight states: GA, IN, LA, MS, NJ, OH, PA, and VA. These results illustrate the marked regional differences of the COVID-19 pandemic in the U.S., with most of the counties with a higher mortality risk concentrated in the North- and South-East regions of the country. Reasons for the increased mortality risk in these areas could be manifold. First, CT and NJ share borders with NY, home to about 5% of worldwide COVID-19 cases^6^. Many people that usually work in NY might reside in CT and NJ border counties which might help the initial spread of the disease. Second, most of these states have higher proportions of at least one minority group (African American and Latino) than the national average. County-average poverty was larger than the national average for GA (20.7 VS 15.6) and LA (22.0 VS 15.6) states. In terms of air pollution, about 50% of states with high mortality risk counties had long-term PM_2.5_ averages above the national average (8.0 µg/m), ranging from 8.1 µg/m (MO) to 11.2 µg/m (DE). Noteworthy, two of the ten most populated counties located in CT and MI were ranked into the 25 most ozone-polluted (Fairfield County, CT), and the 25 most polluted by year-round (Wayne County, MI) across U.S.^40^. Most of the counties with high COVID-19 related mortality risk (71 out of 105) were also counties with lower critical care capacity than national average of 28.4 ICU per 100,000 inhabitants. Our bivariate analysis showed that states like GA, LA, NJ represent most of the counties with high COVID-19 related mortality risk but low healthcare capacity. Those counties had an average of 5.4 ICU per 100,000 in GA, 14.0 per 100,000 in LA, and 16.9 per 100,000 in NJ. With the onset of COVID-19 and the upcoming lift of lockdown measures across U.S., critical healthcare capacity might be potentially overwhelmed in several of these counties not only in ICU beds capacity, but also in mechanical ventilators and staffing. Therefore, counties with high COVID-19 related mortality but low healthcare capacity identified in our study should be prioritized in strategies aimed to diminish the overall number of COVID-19 related deaths including patient relocation, strengthening of critical healthcare infrastructure and supply chains, and staff step-up^41^.

## LIMITATIONS

Our study has several limitations worth noting. First, COVID-19 data that includes comorbidity data are not available at the unit of analysis (county-level). This issue might hamper precise comparisons of the real epidemic burden in specific groups and comorbidities. Second, we analyzed air pollution based on PM_2.5_ measures. Other pollutants including sulfate (SO_4_), nitrate (NO_3_), ammonium (NH_4_), organic matter (OM), black carbon (BC), mineral dust (DUST), and sea-salt (SS) might be needed to produce better pollution estimations. Our result is from 2000 to 2018 and updates with additional a data and analysis warrant in the future. A further limitation relates to the challenges in translating cross-sectional associations into conclusions on causation of COVID-19 related deaths at county-level. Hence, our results should be interpreted with caution.

## CONCLUSIONS

This study is one of the first to explore the population risk determinants of COVID-19 related deaths at a country level, and the use of geospatial approaches to identify vulnerable areas and populations at higher risk of COVID-19 related mortality. These results have significant public health implications to strength the critical healthcare infrastructure for an effective response to the pandemic. The social gradient of health and environment in which most deprived groups are highly vulnerable to more severe health outcomes can be also an important driver of the current geographical and social disparity observed in the current COVID-19 pandemic. Moreover, the substantial regional disparities of the healthcare capacity increase the vulnerability of these areas already at higher risk of COVID-19 related mortality. Therefore, we anticipate that the results from this study can be used to guide the development of strategies for the identification and targeting prevention efforts in these vulnerable high-risk counties with higher proportion of minority groups, poor air quality, and low healthcare capacity.

## Data Availability

The data that support the findings of this study are openly available from the Johns Hopkins University Center for Systems Science and Engineering through a GitHub repository (https://github.com/CSSEGISandData/COVID-19).

## ACKNOWLEDGEMENTS

The authors thank to Centers for Disease Control and Prevention (CDC) for releasing the national surveys in the service of science, and the United States Agencies and other donors supporting these initiatives.

## AUTHOR STATEMENT

Dr Cuadros had full access to all the data in the study and takes responsibility for the integrity of the data and the accuracy of the data analysis. Conceptualization: E.C., M.T., A.H., A.J.B., N.M. D.F.C.; methodology: E.C., M.T., A.H., D.F.C.; software: E.C.; validation: E.C., A.H., D.F.C., A.J.B., N.M.; writing—original draft preparation: E.C., D.F.C., writing—review and editing: E.C., M.T., A.H., A.J.B., N.M.

## PATIENT AND PUBLIC INVOLVEMENT STATEMENT

There were no patients involved in this research.

## ETHICS APPROVAL STATEMENT

The data that support the findings of this study are openly available from the Johns Hopkins University Center for Systems Science and Engineering through a GitHub repository (https://github.com/CSSEGISandData/COVID-19). Therefore, our study did not require an ethics approval because there were no human participants involved in this research, and it relies entirely on previously published data.

## SUPLEMENTARY MATERIALS

## APPENDIX A: Covariate selection criteria and definitions

All covariates were selected according to an evidence synthesis process of relevant references ^2,11-15^. This section describes the sources of sociodemographic, concomitant comorbidities, environment and the spatial explicit cofactors according to the implied graph.

**Figure S1.**
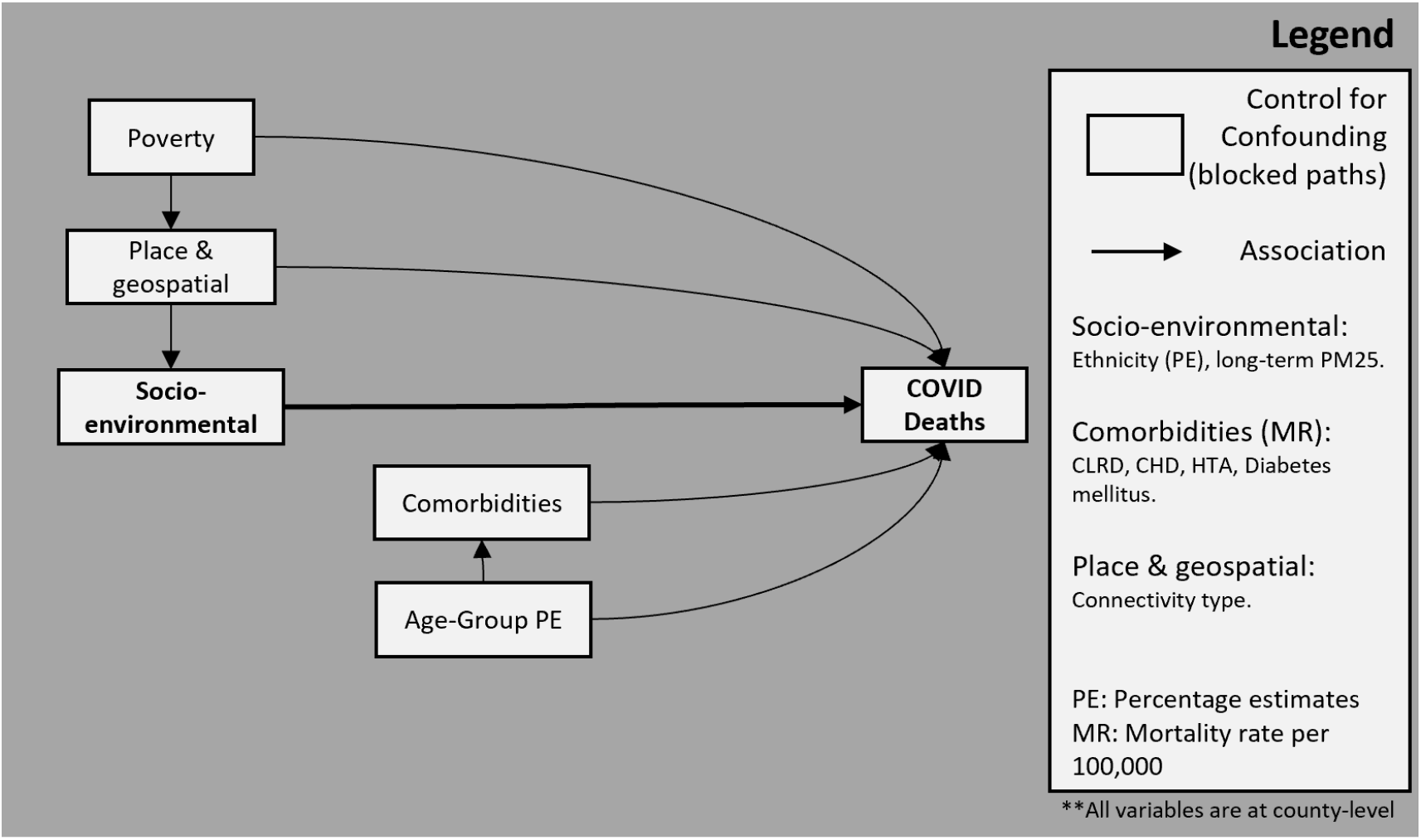
Hypothetical DAG for the study

All covariates ACS surveys are readily available and are described as follows:

Age: Percent estimate of total population according to the following groups: under 25 years, 25 to 34 years, 35-44 years, 45 to 59 years, 60 to 74 years, over 75 years. Variable names: DP05_0005PE, DP05_0006PE, DP05_0007PE, DP05_0008PE, DP05_0009PE, DP05_0010PE, DP05_0012PE, DP05_0013PE, DP05_0014PE, DP05_0015PE, DP05_0016PE, DP05_0017PE.

Poverty: According to the census bureau, the income money threshold and the consumer Price Index (CPI-U). If a family’s total income is less than the family’s threshold, then every individual of that family is considered in poverty^42^. Variable name: S0601_C01_049E.

Race: Self-identification of a person with one or more social groups. Percent estimate of white, black. Variable names: DP05_0037PE, DP05_0038PE.

Ethnicity: Hispanic origin or not. Hispanics may report as any race. Percent estimate of Latino population. Variable names: DP05_0071P.

Underlying cause of death: Four COVID-related underlying cause of death including Chronic lower respiratory diseases (ICD-10: J40-J47), diabetes mellitus (ICD-10: E10-E14), hypertensive diseases (ICD-10: I10-I15), and ischemic heart diseases (ICD-10: I20-I25) were extracted from the CDC Wonder database^9^ using the ICD-10 standard code.

PM2.5: For the exposure estimates, PM2.5 cross-validated exposure estimates were produced by van Donekelaar et al^10^.

**Figure S2.**
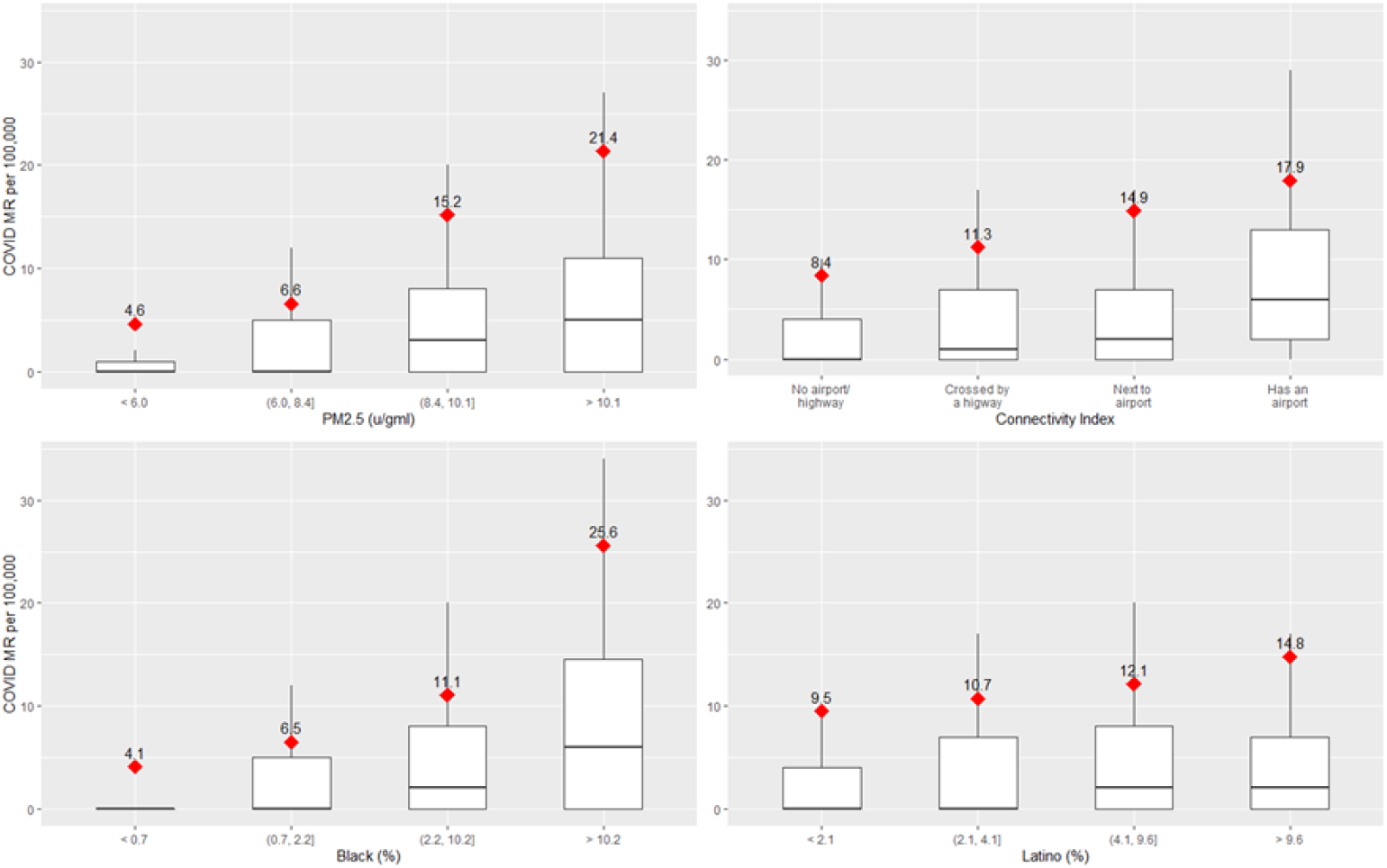
Exploratory data analysis of MR in PM2.5 quantiles, connectivity index, black and latino population quantiles.

**Table S1.**
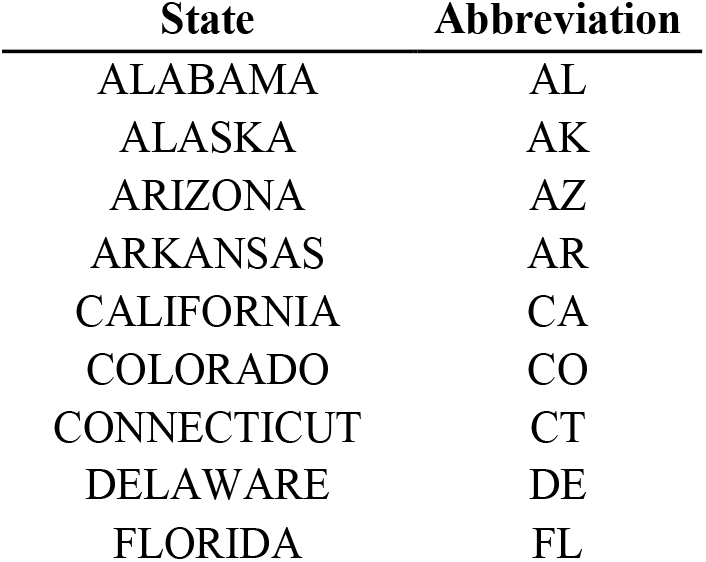
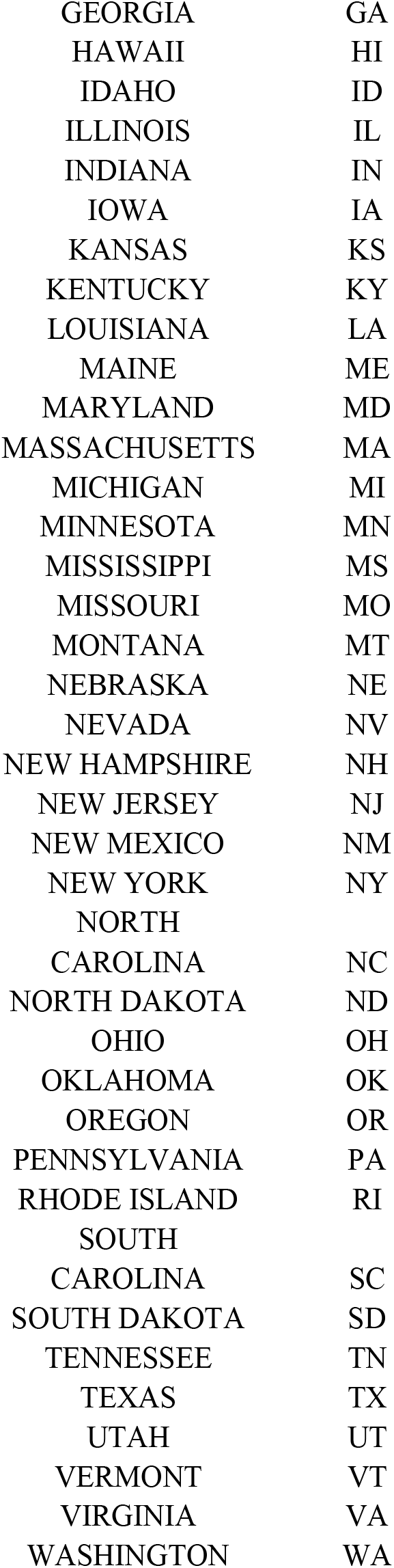
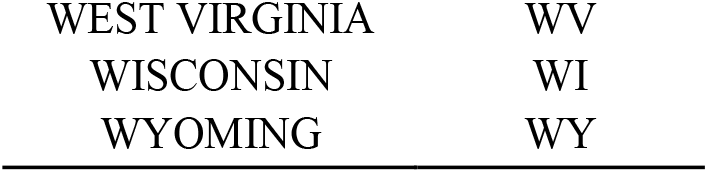
State Abbreviations List.

| State | Abbreviation |
| --- | --- |
| ALABAMA | AL |
| ALASKA | AK |
| ARIZONA | AZ |
| ARKANSAS | AR |
| CALIFORNIA | CA |
| COLORADO | CO |
| CONNECTICUT | CT |
| DELAWARE | DE |
| FLORIDA | FL |
| GEORGIA | GA |
| HAWAII | HI |
| IDAHO | ID |
| ILLINOIS | IL |
| INDIANA | IN |
| IOWA | IA |
| KANSAS | KS |
| KENTUCKY | KY |
| LOUISIANA | LA |
| MAINE | ME |
| MARYLAND | MD |
| MASSACHUSETTS | MA |
| MICHIGAN | MI |
| MINNESOTA | MN |
| MISSISSIPPI | MS |
| MISSOURI | MO |
| MONTANA | MT |
| NEBRASKA | NE |
| NEVADA | NV |
| NEW HAMPSHIRE | NH |
| NEW JERSEY | NJ |
| NEW MEXICO | NM |
| NEW YORK | NY |
| NORTH |  |
| CAROLINA | NC |
| NORTH DAKOTA | ND |
| OHIO | OH |
| OKLAHOMA | OK |
| OREGON | OR |
| PENNSYLVANIA | PA |
| RHODE ISLAND | RI |
| SOUTH |  |
| CAROLINA | SC |
| SOUTH DAKOTA | SD |
| TENNESSEE | TN |
| TEXAS | TX |
| UTAH | UT |
| VERMONT | VT |
| VIRGINIA | VA |
| WASHINGTON | WA |
| WEST VIRGINIA | WV |
| WISCONSIN | WI |
| WYOMING | WY |

## APPENDIX B: Statistical Analysis

In certainly contexts where data belongs to certain group or cluster (nested structure), multilevel level models (MLMs) are a recommended approach to assess the group effects19. For this case study in which each state context and policies can influence health outcomes, MLMs offer a great flexibility to understand COVID-related mortality by avoiding overall averaging and retaining variation across subsamples (counties) on different states modeled as random intercepts. Therefore, we used a Bayesian multilevel analysis to assess the risk of COVID-related death per county adjusted by the selected covariates. For the model selection, complementarily to the evidence synthesis, we evaluated each variable after adjusting by age. All variables who have an RHAT less than 1.05 and acceptable effective sample size were allowed to be included in the final adjusted model.

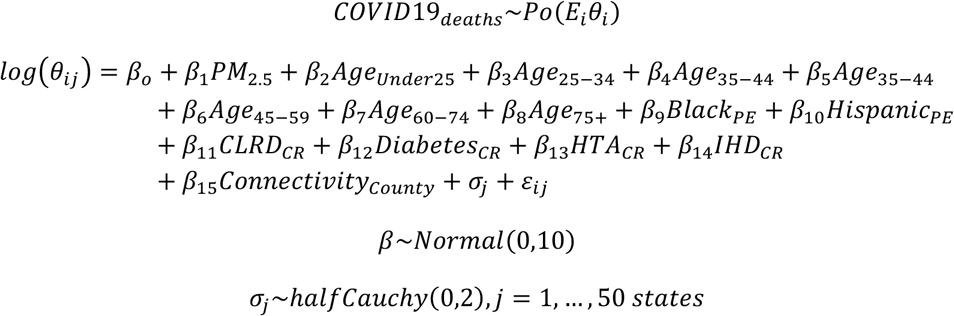

**Table S2.**
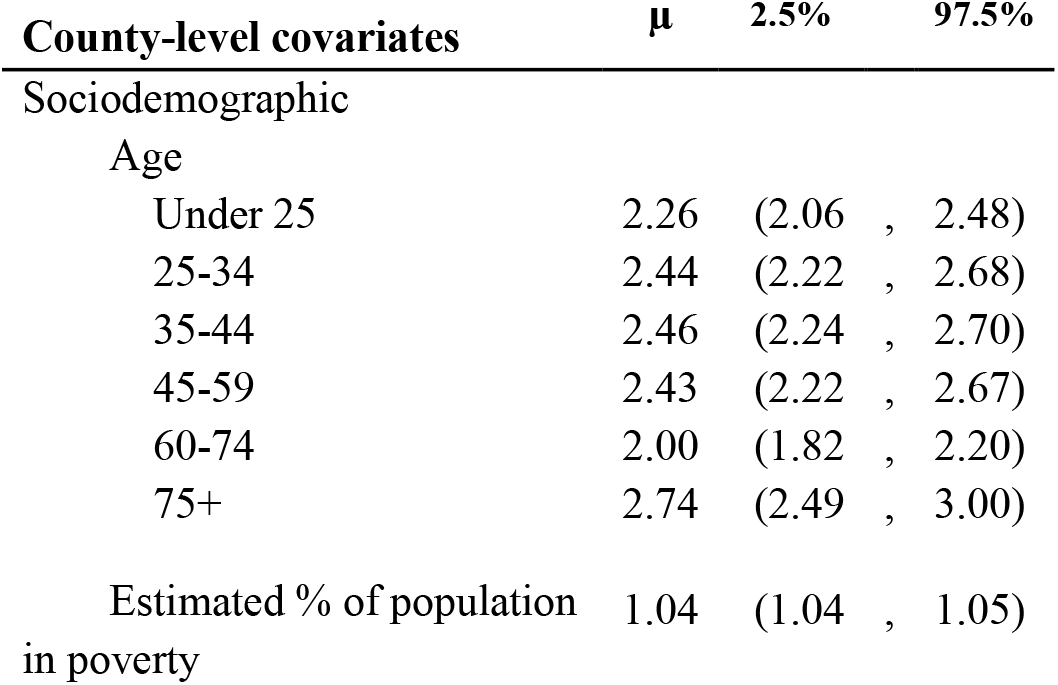
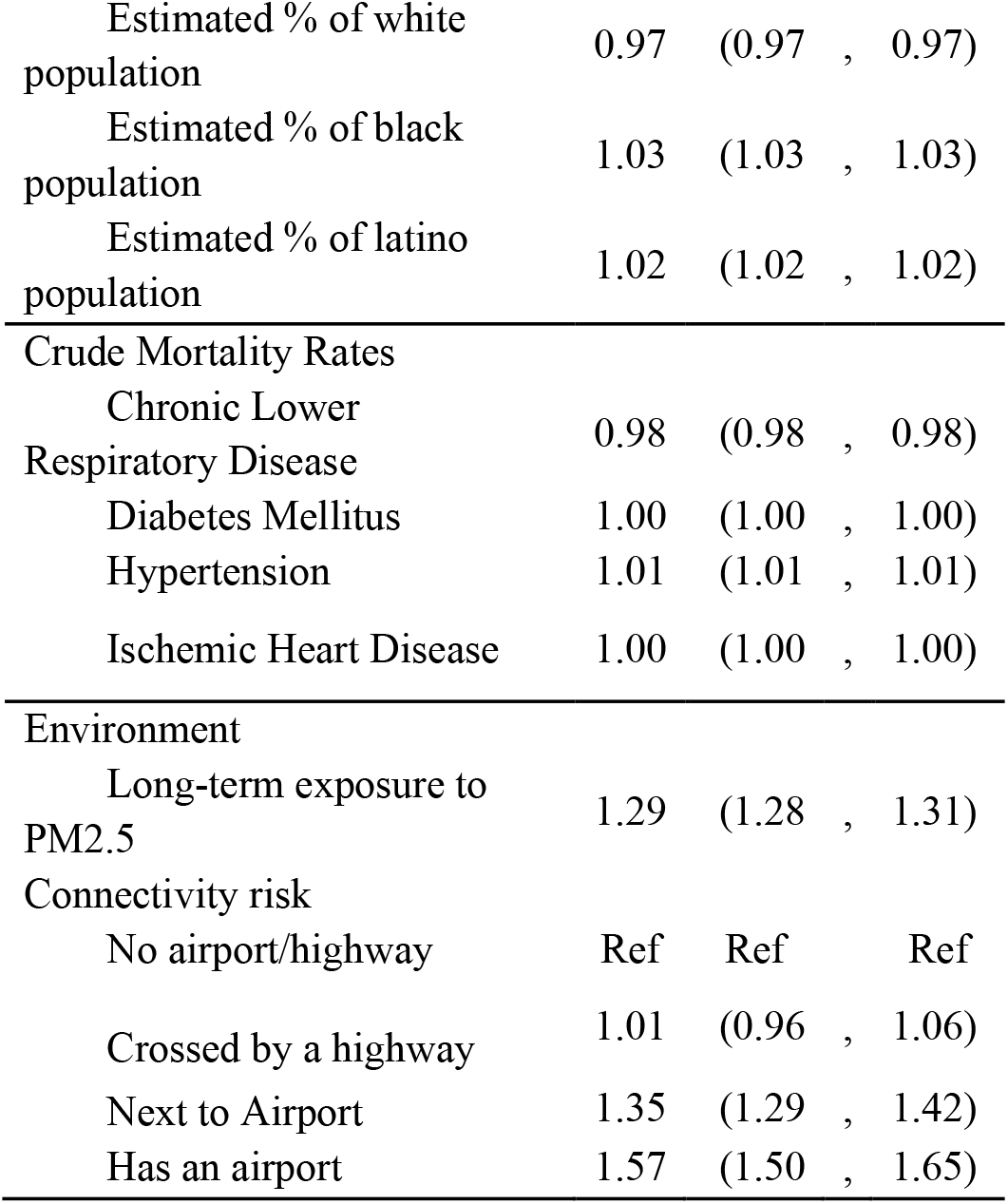
Unadjusted county-level predictors of COVID19 deaths.

### Convergence diagnostics

Different converge diagnostics were used to warrant the reproducibility and the right results on all Markov chain Monte Carlo models.

All included covariates had an R-hat < 1.01. If chains have not mixed well (ie, the between- and within-chain estimates do not agree), R-hat is larger than 1.

The effective sample size (ESS) captures how many independent draws contain the same amount of information as the dependent sample obtained by the NUTS sampler. We recommend a minimum ESS greater than 100 times the number of chains (4).

**Figure S3.**
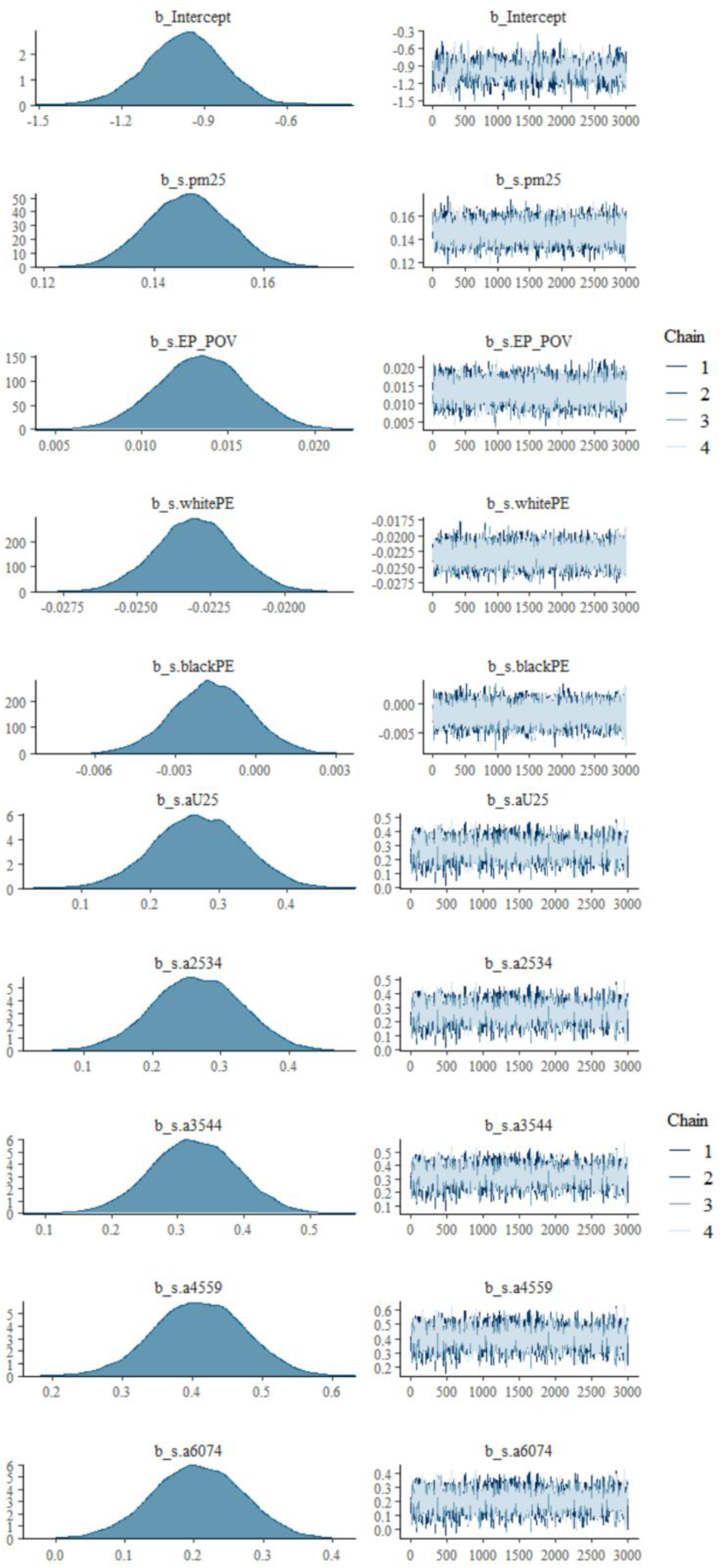
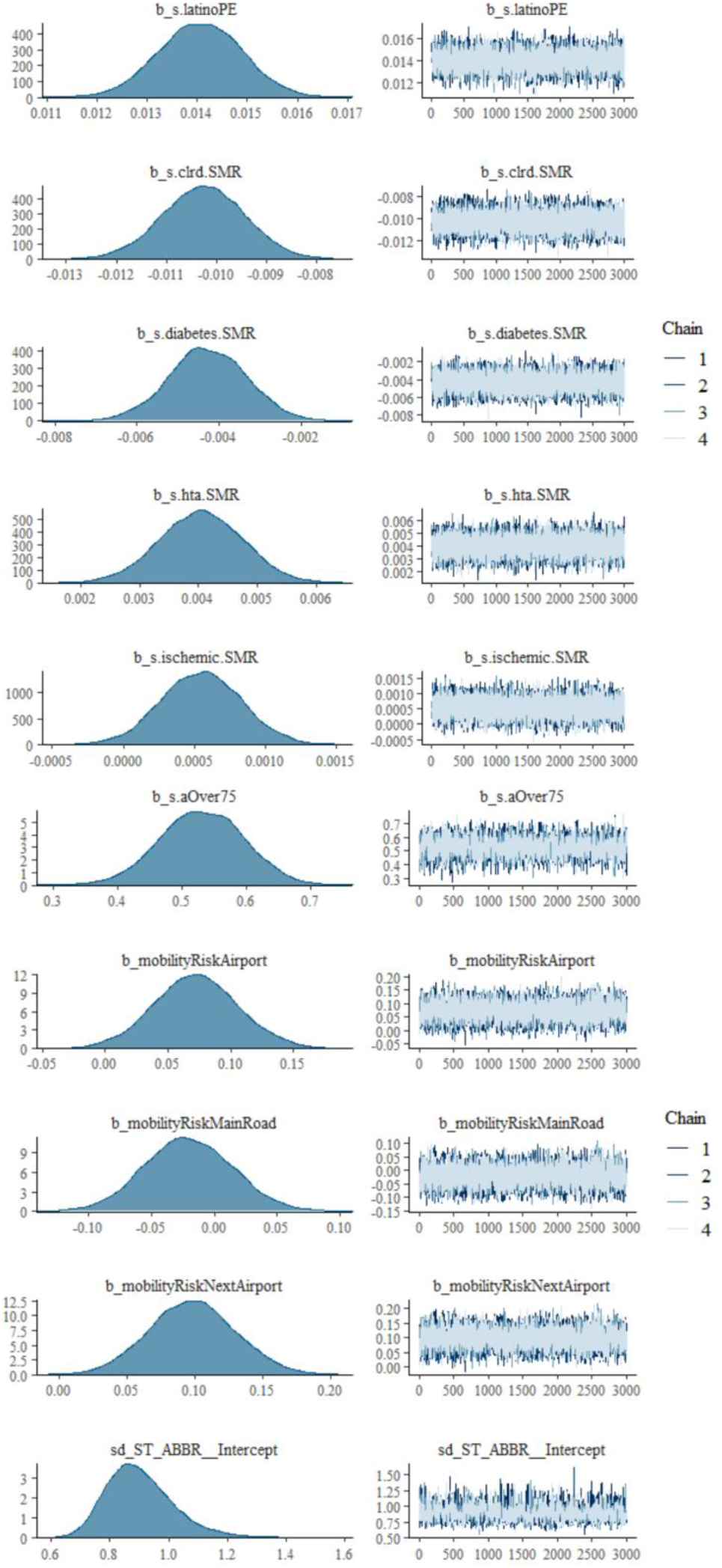
MCMC sampling confirming the convergence properties of the model.

**Figure S4.**
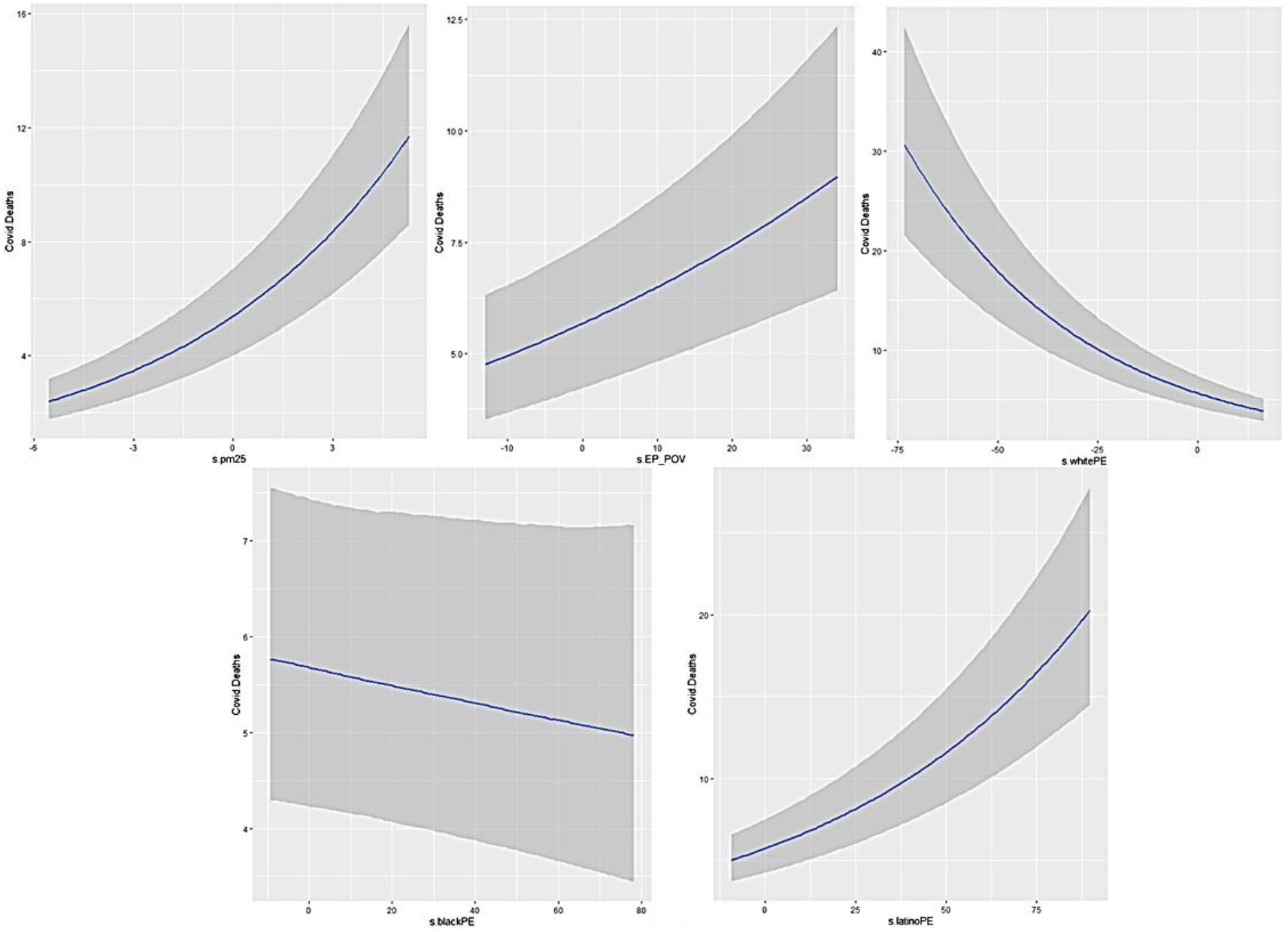
Marginal effects showing each significant variable’s effect when the others are set to a value of zero.

## APPENDIX C: Disease mapping

Disease mapping is the process of visually depicting geographically indexed data in a spatial referenced distribution for explanatory purposes. Small area disease models are commonly used to quantify risk factors using lattice data.

The general model in Small area disease models is expressed as

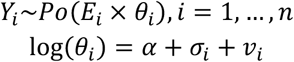

Where *α* denotes the overall risk level, *σ*_*i*_ is the random effect that models the state-level dependence of counties’ relative risk, and *v*_*i*_ is the uncorrelated variance.

**Table S3.**
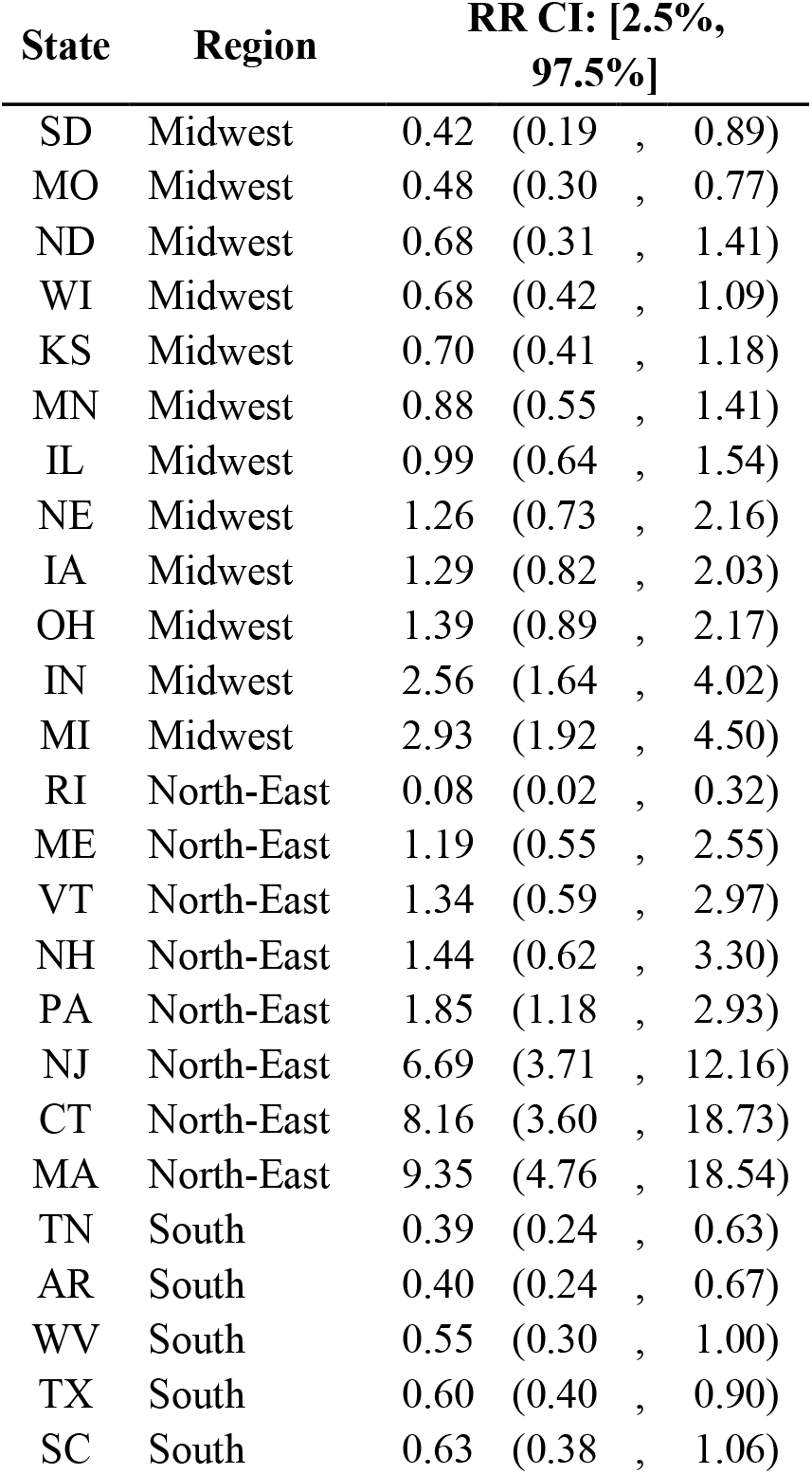
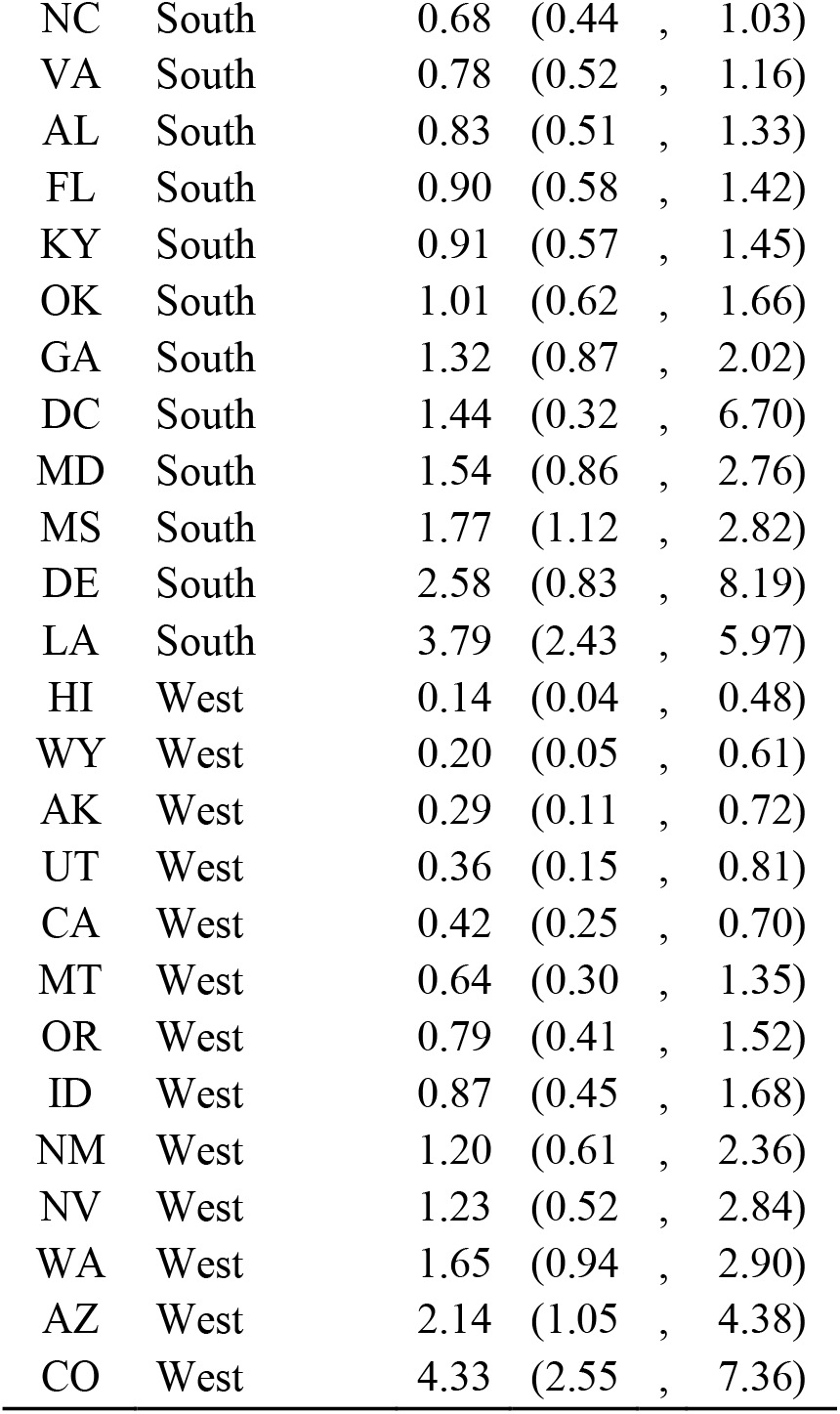
Relative risk by state.

| State | Region | RR CI: [2.5%,<br>97.5%] |  |
| --- | --- | --- | --- |
| SD | Midwest | 0.42 | (0.19 , 0.89) |
| MO | Midwest | 0.48 | (0.30 , 0.77) |
| ND | Midwest | 0.68 | (0.31 , 1.41) |
| WI | Midwest | 0.68 | (0.42 , 1.09) |
| KS | Midwest | 0.70 | (0.41 , 1.18) |
| MN | Midwest | 0.88 | (0.55 , 1.41) |
| IL | Midwest | 0.99 | (0.64 , 1.54) |
| NE | Midwest | 1.26 | (0.73 , 2.16) |
| IA | Midwest | 1.29 | (0.82 , 2.03) |
| OH | Midwest | 1.39 | (0.89 , 2.17) |
| IN | Midwest | 2.56 | (1.64 , 4.02) |
| MI | Midwest | 2.93 | (1.92 , 4.50) |
| RI | North-East | 0.08 | (0.02 , 0.32) |
| ME | North-East | 1.19 | (0.55 , 2.55) |
| VT | North-East | 1.34 | (0.59 , 2.97) |
| NH | North-East | 1.44 | (0.62 , 3.30) |
| PA | North-East | 1.85 | (1.18 , 2.93) |
| NJ | North-East | 6.69 | (3.71 , 12.16) |
| CT | North-East | 8.16 | (3.60 , 18.73) |
| MA | North-East | 9.35 | (4.76 , 18.54) |
| TN | South | 0.39 | (0.24 , 0.63) |
| AR | South | 0.40 | (0.24 , 0.67) |
| WV | South | 0.55 | (0.30 , 1.00) |
| TX | South | 0.60 | (0.40 , 0.90) |
| SC | South | 0.63 | (0.38 , 1.06) |
| NC | South | 0.68 | (0.44 , 1.03) |
| VA | South | 0.78 | (0.52 , 1.16) |
| AL | South | 0.83 | (0.51 , 1.33) |
| FL | South | 0.90 | (0.58 , 1.42) |
| KY | South | 0.91 | (0.57 , 1.45) |
| OK | South | 1.01 | (0.62 , 1.66) |
| GA | South | 1.32 | (0.87 , 2.02) |
| DC | South | 1.44 | (0.32 , 6.70) |
| MD | South | 1.54 | (0.86 , 2.76) |
| MS | South | 1.77 | (1.12 , 2.82) |
| DE | South | 2.58 | (0.83 , 8.19) |
| LA | South | 3.79 | (2.43 , 5.97) |
| HI | West | 0.14 | (0.04 , 0.48) |
| WY | West | 0.20 | (0.05 , 0.61) |
| AK | West | 0.29 | (0.11 , 0.72) |
| UT | West | 0.36 | (0.15 , 0.81) |
| CA | West | 0.42 | (0.25 , 0.70) |
| MT | West | 0.64 | (0.30 , 1.35) |
| OR | West | 0.79 | (0.41 , 1.52) |
| ID | West | 0.87 | (0.45 , 1.68) |
| NM | West | 1.20 | (0.61 , 2.36) |
| NV | West | 1.23 | (0.52 , 2.84) |
| WA | West | 1.65 | (0.94 , 2.90) |
| AZ | West | 2.14 | (1.05 , 4.38) |
| CO | West | 4.33 | (2.55 , 7.36) |

**Figure S5.**
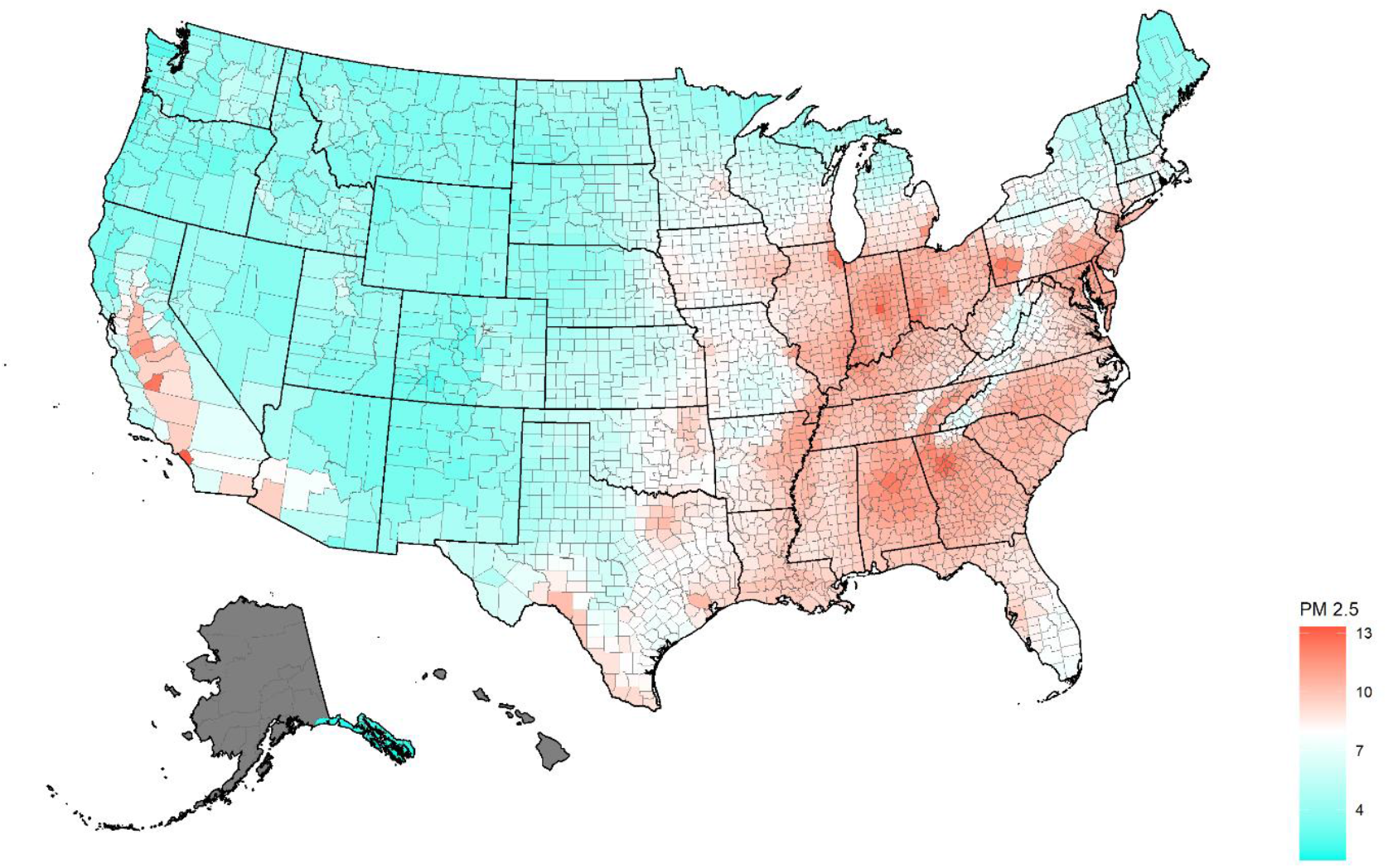
U.S. 2000 to 2018 Long-Term Mean PM2.5 Concentrations by County, mean=7.98 µg/m (range is 1.42-13.30).

**Figure S6.**
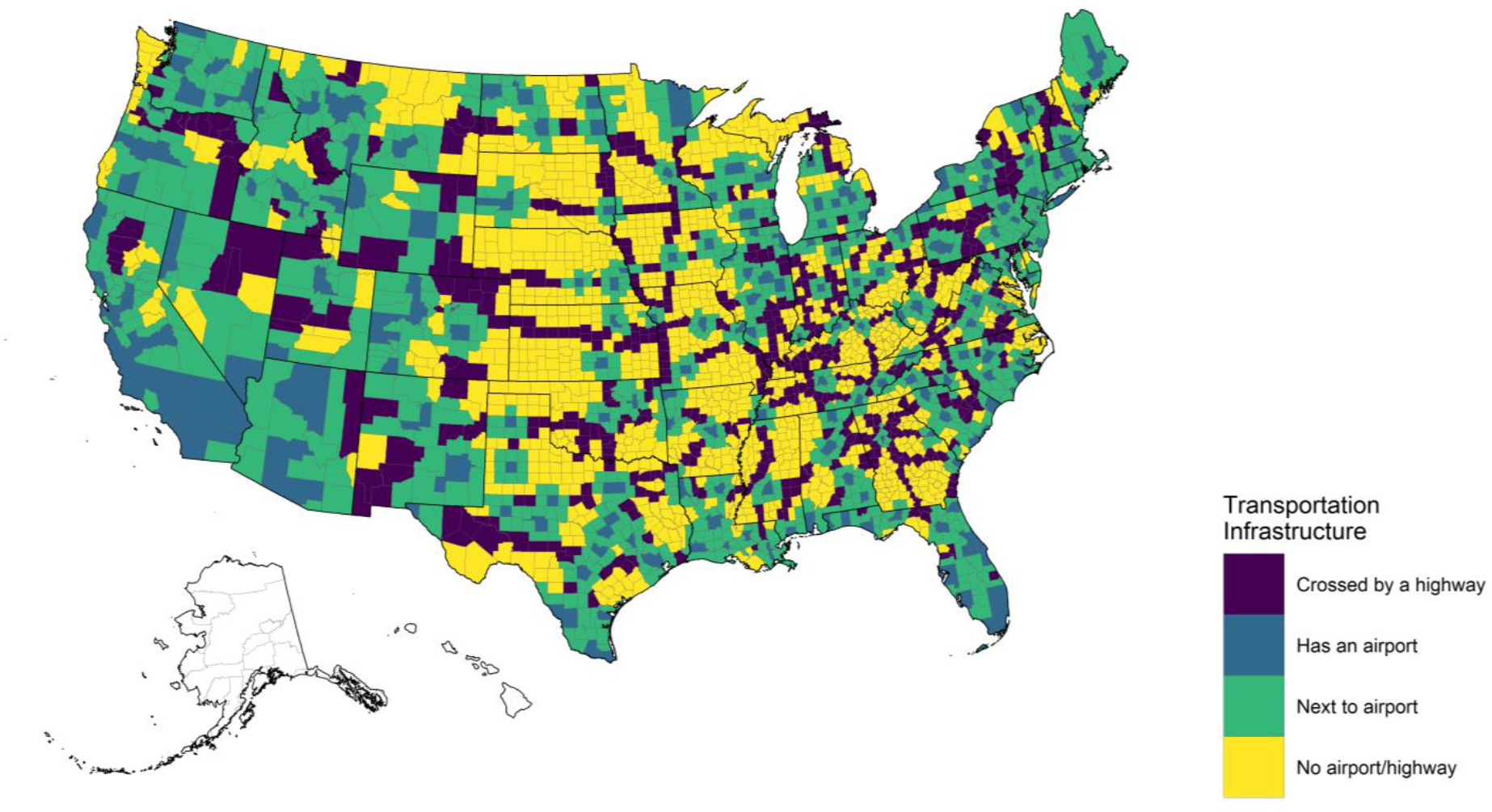
U.S. Connectivity index by county.

## REFERENCES

1. Richardson S, Hirsch JS, Narasimhan M, et al. Presenting Characteristics, Comorbidities, and Outcomes Among 5700 Patients Hospitalized With COVID-19 in the New York City Area. JAMA. 2020.

2. Williamson E, Walker AJ, Bhaskaran KJ, et al. OpenSAFELY: factors associated with COVID-19-related hospital death in the linked electronic health records of 17 million adult NHS patients. medRxiv. 2020:2020.2005.2006.20092999.

3. Remuzzi A, Remuzzi G. COVID-19 and Italy: what next? The Lancet. 2020;395(10231):1225–1228.

4. Li R, Rivers C, Tan Q, Murray MB, Toner E, Lipsitch M. Estimated Demand for US Hospital Inpatient and Intensive Care Unit Beds for Patients With COVID-19 Based on Comparisons With Wuhan and Guangzhou, China. JAMA Network Open. 2020;3(5):e208297–e208297.

5. University JH. 2019 Novel Coronavirus COVID-19 (2019-nCoV) Data Repository by Johns Hopkins CSSE. https://github.com/CSSEGISandData/COVID-19. accessed 06/07/2020, 2020.

6. Nature. Coronavirus: the first three months as it happened. 2020; https://www.nature.com/articles/d41586-020-00154-w. Accessed 05/19/2020.

7. Bureau USC. American Community Survey 2014-2018 5-Year Estimates. Retrieved from https://data.census.gov/cedsci/table?g=0100000US.0050000&tid=ACSST0100005Y0102018.S0100101&hidePreview=false&vintage=0102018&layer=VT_0102018_0100050_0100000_PY_D0100001&cid=DP0100005_0100001E. Available at. Accessed 04/20/2020.

8. Gregory EW, Hallisey E, Heitgerd JL, Lewis B. A Social Vulnerability Index for Disaster Management. Paper presented at: Journal of Homeland Security and Emergency Management 2011.

9. Centers for Disease Control and Prevention NCfHS. Underlying Cause of Death 1999-2018 on CDC WONDER Online Database, released in 2020. Data are from the Multiple Cause of Death Files, 1999-2018, as compiled from data provided by the 1957 vital statistics jurisdictions through the Vital Statistics Cooperative Program. Available at: http://wonder.cdc.gov/ucd-icd10.html. Accessed Apr 26, 2020.

10. van Donkelaar A, Martin RV, Li C, Burnett RT. Regional Estimates of Chemical Composition of Fine Particulate Matter Using a Combined Geoscience-Statistical Method with Information from Satellites, Models, and Monitors. Environmental Science & Technology. 2019;53(5):2595–2611.

11. Niedzwiedz CL, Donnell CA, Jani BD, et al. Ethnic and socioeconomic differences in SARS-CoV-2 infection: prospective cohort study using UK Biobank. medRxiv. 2020:2020.2004.2022.20075663.

12. Halpin DMG, Faner R, Sibila O, Badia JR, Agusti A. Do chronic respiratory diseases or their treatment affect the risk of SARS-CoV-2 infection? The Lancet Respiratory Medicine. 2020;8(5):436–438.

13. Pansini R, Fornacca D. Initial evidence of higher morbidity and mortality due to SARS-CoV-2 in regions with lower air quality. medRxiv. 2020:2020.2004.2004.20053595.

14. Fattorini D, Regoli F. Role of the atmospheric pollution in the Covid-19 outbreak risk in Italy. medRxiv. 2020:2020.2004.2023.20076455.

15. Team CC-R. Preliminary Estimates of the Prevalence of Selected Underlying Health Conditions Among Patients with Coronavirus Disease 2019. 2020;69(13):382–386.

16. Hernán MA, Hernández-Díaz S, Robins JM. A Structural Approach to Selection Bias. Epidemiology. 2004;15(5).

17. Bureau USC. Race & Ethnicity. https://www.census.gov/mso/www/training/pdf/race-ethnicity-onepager.pdf. Accessed 04/20/2020.

18. von Elm E, Altman DG, Egger M, Pocock SJ, Gotzche PC, Vandenbroucke JP. The Strengthening the Reporting of Observational Studies in Epidemiology (STROBE) Statement: Guidelines for Reporting Observational Studies. Epidemiology. 2007;18(6):800–804.

19. Gelman A. Prior distributions for variance parameters in hierarchical models (comment on article by Browne and Draper). Bayesian Anal. 2006;1(3):515–534.

20. Hoffman MD, Gelman A. he No-U-Turn Sampler: Adaptively Setting Path Lengths in Hamiltonian Monte Carlo. Journal of Machine Learning Research. 2014;15(47):1593–1623.

21. Healthcare D. USA Hospital Beds. 2020; https://coronavirus-resources.esri.com/datasets/1044bb19da8d4dbfb6a96eb1b4ebf629_0?geometry=80.507%2C-16.820%2C-105.469%2C72.123. Accessed 05/05/2020.

22. raster: Geographic Data Analysis and Modeling [computer program]. 2019.

23. R: A Language and Environment for Statistical Computing [computer program]. Version 3.5.2. Vienna, Austria: R Foundation for Statistical Computing; 2018.

24. Bürkner P-C. brms: An R Package for Bayesian Multilevel Models Using Stan. Journal of Statistical Software; Vol 1, Issue 1 (2017). 2017.

25. SpatialEpi: Methods and Data for Spatial Epidemiology [computer program]. Version R package version 1.2.32018.

26. Owen WF, Jr., Carmona R, Pomeroy C. Failing Another National Stress Test on Health Disparities. JAMA. 2020.

27. Hygiene NDoHaM. COVID-19: Data. 2020; https://www1.nyc.gov/site/doh/covid/covid-19-data.page. Accessed 05/17/2020.

28. Hernandez A, Correa-Agudelo E, Kim H, et al. On the impact of early non-pharmaceutical interventions as containment strategies against the COVID-19 pandemic. medRxiv. 2020.

29. Kelly FJ, Fussell JC. Air pollution and airway disease. Clinical and experimental allergy : journal of the British Society for Allergy and Clinical Immunology. 2011;41(8):1059–1071.

30. Wu S, Ni Y, Li H, et al. Short-term exposure to high ambient air pollution increases airway inflammation and respiratory symptoms in chronic obstructive pulmonary disease patients in Beijing, China. Environment international. 2016;94:76–82.

31. Minelli C, Wei I, Sagoo G, Jarvis D, Shaheen S, Burney P. Interactive effects of antioxidant genes and air pollution on respiratory function and airway disease: a HuGE review. American journal of epidemiology. 2011;173(6):603–620.

32. Viegi G, Simoni M, Scognamiglio A, et al. Indoor air pollution and airway disease. The international journal of tuberculosis and lung disease : the official journal of the International Union against Tuberculosis and Lung Disease. 2004;8(12):1401–1415.

33. Ulmer WT. [Air pollution and airway disease]. Der Internist. 1985;26(4):233–240.

34. Karakatsani A, Analitis A, Perifanou D, et al. Particulate matter air pollution and respiratory symptoms in individuals having either asthma or chronic obstructive pulmonary disease: a European multicentre panel study. Environmental health : a global access science source. 2012;11:75.

35. Vineis P, Hoek G, Krzyzanowski M, et al. Air pollution and risk of lung cancer in a prospective study in Europe. International journal of cancer. 2006;119(1):169–174.

36. Raaschou-Nielsen O, Vineis P, Brunekreef B, et al. Air pollution and lung cancer in Europe - authors’ reply. The Lancet Oncology. 2013;14(11):e440.

37. Sax SN, Zu K, Goodman JE. Air pollution and lung cancer in Europe. The Lancet Oncology. 2013;14(11):e439–440.

38. Nawrot TS, Nackaerts K, Hoet PH, Nemery B. Lung cancer mortality and fine particulate air pollution in Europe. International journal of cancer. 2007;120(8):1825–1826; author reply 1827.

39. Richardson S, Hirsch JS, Narasimhan M, et al. Presenting Characteristics, Comorbidities, and Outcomes Among 5700 Patients Hospitalized With COVID-19 in the New York City Area. JAMA. 2020.

40. American Lung Association. State of the Air 2020. American Lung Association; 2020.

41. Institute for Health Metrics and Evaluation. New COVID-19 forecasts: US hospitals could be overwhelmed in the second week of April by demand for ICU beds, and US deaths could total 81,000 by July. 2020; www.healthdata.org/news-release/new-covid-19-forecasts-us-hospitals-could-be-overwhelmed-second-week-april-demand-icu. Accessed 05/10/2020.

42. Bureau C. How the Census Bureau Measures Poverty. https://www.census.gov/topics/income-poverty/poverty/guidance/poverty-measures.html. Accessed 05/11/2020.

